# Prediction and diagnosis of chronic kidney disease development and progression using machine-learning: protocol for a systematic review and meta-analysis of reporting standards and model performance

**DOI:** 10.1101/2022.11.24.22282661

**Authors:** Fangyue Chen, Piyawat Kantagowit, Tanawin Nopsopon, Arisa Chuklin, Krit Pongpirul

## Abstract

Chronic Kidney disease (CKD) is an important yet under-recognized contributor to morbidity and mortality globally. Machine-learning (ML) based decision support tools have been developed across many aspects of CKD care. Notably, algorithms developed in the prediction and diagnosis of CKD development and progression may help to facilitate early disease prevention, assist with early planning of renal replacement therapy, and offer potential clinical and economic benefits to patients and health systems. Clinical implementation can be affected by the uncertainty surrounding the methodological rigor and performance of ML-based models. This systematic review aims to evaluate the application of prognostic and diagnostic ML tools in CKD development and progression.

The protocol has been prepared using the Preferred Items for Systematic Review and Meta-analysis Protocols (PRISMA-P) guidelines. The systematic review protocol for CKD prediction and diagnosis have been registered with the International Prospective Register of Systematic Reviews (PROSPERO) (CRD42022356704, CRD42022372378). A systematic search will be undertaken of PubMed, Embase, the Cochrane Central Register of Controlled Trials (CENTRAL), the Web of Science, and the IEEE Xplore digital library. Studies in which ML has been applied to predict and diagnose CKD development and progression will be included. The primary outcome will be the comparison of the performance of ML-based models with non-ML-based models. Secondary analysis will consist of model use cases, model construct, and model reporting quality.

This systematic review will offer valuable insight into the performance and reporting quality of ML-based models in CKD diagnosis and prediction. This will inform clinicians and technical specialists of the current development of ML in CKD care, as well as direct future model development and standardization.

## Introduction

Chronic kidney disease (CKD) is an important non-communicable disease that contributes to significant morbidity and mortality on a global scale, directly or through cardiovascular diseases attributable to impaired kidney function. Its estimated prevalence ranges from 9.1 to 15.1%. It has increased by 29.3% since 1990 due to the increase in the major chronic diseases that contribute to its development, notably diabetes mellitus and hypertension (1,2). Nevertheless, CKD is underrecognized by patients, clinicians, and health authorities. The disease often progresses insidiously to end-stage kidney disease (ESKD) with late presentation of symptoms and signs (3,4). The life-sustaining treatment for ESKD, renal replacement therapy, poses a significant economic burden on patients and health systems, meaning that currently, an estimated 47 to 73% of individuals are unable to receive it, leaving around 2.3 million individuals dying prematurely (5). Strategies to prevent or delay CKD onset and progression can potentially lower overall morbidity and mortality while minimizing cost.

Machine Learning (ML), a subset of artificial intelligence (AI), has seen exponential growth across healthcare (6,7). ML utilizes a specific dataset to generate an algorithm that employs unknown or varied combinations of complex features and weights to predict the outcome of future inputs (8). ML-based decision support tools have been developed across many aspects of CKD care across disease prevention, diagnosis, and treatment (7), fuelled by the growth in volume and variety of big data in nephrology and healthcare in general (9,10). Notably, algorithms developed in the prediction and diagnosis of CKD development and progression to ESKD may help to facilitate early disease prevention, assist with early care planning, and allocate resources for the most significant clinical benefit (11-15).

Despite the growing promise of ML, several factors can hinder its clinical uptake. These include uncertainty surrounding the performance of ML and the methodological rigor behind its development. Non-ML-based prediction tools for CKD progression and prognosis have been developed and validated, such as the Kidney Failure Risk Equation, which has been used clinically to guide referrals to multidisciplinary CKD clinics (16-18). Comparisons have been made between ML and non-ML-based prediction tools in general, specifically to chronic diseases and prediction of acute kidney injury, which found similar performance between prediction models developed with ML and conventional logistic regression (LR) techniques (19-21). In addition, previous studies have questioned the reporting quality and methodology of CKD prediction models (22,23), as well as other AI-based models in imaging (24), oncology (25), and COVID-19 (26).

This systematic review aims to provide a comprehensive, in-depth summary and evaluation of ML-based diagnostic and prognostic tools for CKD development and progression, which will help to better direct future research strategy and methodology in developing ML algorithms in CKD care.

The proposed systematic review aims to answer the following questions:

1. How do ML-based prediction tools in CKD development and progression perform compared with tools developed using conventional techniques?
2. What are the use cases and constructs of these prediction tools?
3. How are the methodological characteristics and reporting quality of the ML-based tools?

## Materials and Methods

The systematic review protocol was registered with the International Prospective Register of Systematic Reviews (PROSPERO) on 26/09/22 for CKD prediction (CRD42022356704) and CKD diagnosis (CRD42022372378). The protocol followed the Preferred Reporting Items for Systematic Review and Meta-Analysis Protocols (PRISMA-P) 2015 statement (27). The Checklist for critical Appraisal and data extraction for systematic Reviews of prediction Modelling Studies (CHARMS) has been used to formulate review questions and data extraction (28).

### Study eligibility criteria

#### Study designs

Any peer-reviewed primary studies which assessed a prediction algorithm that utilizes ML techniques applied to clinical problems in the prediction and diagnosis of chronic kidney disease development and progression, including those for CKD screening, CKD prevention, profiling of biomarkers contributing towards CKD, profiling of risk factors leading to CKD, estimation of glomerular filtration rate (GFR) and creatinine levels, prediction of occurrence of CKD, prediction of CKD stages, CKD diagnosis, CKD prognostication, prediction of CKD progression to ESKD and/or requirement for renal replacement therapy, ESKD diagnosis will be included.

Exclusion criteria are 1) Studies that utilize only image-based inputs as the different model development processes require alternative extraction and appraisal tools; 2) studies assessing prediction models of CKD complications other than its progression, including non-exhaustively anemia, electrolyte disturbances, bone disorders, and cardiovascular events; 3) prediction model of RRT including non-exhaustively the choice of RRT modalities which are hemodialysis, peritoneal dialysis, and renal transplantation, and 4) studies reporting only treatment-related outcomes of CKD such as adverse events, rate of complications, the management of complications; 5) informal publication types such as case studies, commentaries, letters to the editor, editorials, meeting abstracts, proceeding papers, conference abstracts, protocols, guidelines, and recommendations; 6) review articles such as narrative review, overview, systematic review, meta-analysis; 7) animal studies.

#### Study participants

Adult humans whose age was equal to or more than 18 years old.

#### Types of interventions

The studies will present prediction models utilizing ML techniques, including non-exhaustively various regression techniques, decision trees, random forests, support vector machines, K-nearest neighbor, and neural networks, as defined by individual studies. The models will be for the prediction and diagnosis of chronic kidney disease development and progression with or without mention of ESKD.

#### Comparators

We will include studies that compare the performance of ML-based prediction models with those that utilize conventional techniques, including non-exhaustively those that use logistic regression (including penalized LR), cox regression, Poisson regression, least squares linear separation, generalized additive models, discriminant analysis, generalized estimation equations, risk scores, and expert views. Studies that utilize only ML-based tools will also be included.

### Study outcomes

#### Primary outcome

- Performance comparison of ML-based and non-ML-based prediction tools in CKD development and progression

#### Secondary outcomes

- ML-based model use case
- Performance of ML-based prediction tool in CKD development and progression
- Stages of model development (internal or external validation or clinical implementation)
- Model development team specialty and the involvement of model end-user such as clinicians during model development
- Evidence of model reporting quality description
- Characteristics of the dataset (size of training, validation and testing datasets, source of dataset, population group, data period, length of follow-up)
- Prediction model construct including ML-based and non-ML based techniques
- Predictor characteristics and selection
- Model outcome characteristics and selection
- Model performance measures used

### Information sources and search strategy

We will search through five databases: PubMed, Embase, the Cochrane Central Register of Controlled Trials (CENTRAL), Web of Science, and the IEEE Xplore digital library. The search strategy is constructed by two health information specialists with systematic review experiences, combining search terms and subject headings (MeSH) related to “machine learning,” “artificial intelligence,” “chronic kidney disease,” and “End-stage kidney disease” **(See Additional File)**. PubMed, Embase (OVID interface, 1947 onwards), Web of Science, and CENTRAL were chosen for their broad coverage across biomedical, nursing, allied health, and general scientific literature, while IEEE Xplore was included for coverage of more technical literature in data science. Additional articles will be retrieved by manually scrutinizing the reference lists of relevant publications.

### Study records

#### Data management

Following database searching, studies will be populated into Covidence systematic review software (29), which will manage study selection and data extraction.

#### Selection process

We will carry out two stages of screening. After study de-duplication through Covidence, two reviewers will screen the titles and abstracts of potential studies independently. We will eliminate abstracts in the initial screen if they do not report ML-based prediction models in CKD.

Included studies will undergo full-text review against the full eligibility criteria. Reasons for exclusion will be recorded for each study. Disagreement between two reviewers at each article screening and selection stage will be resolved by consensus and a third person if necessary. The PRISMA 2020 flow diagram will be generated to describe the workflow and identification of included studies for the systematic review (30).

#### Data collection and management

The data extraction form will be designed prospectively before data collection and will be pilot tested and refined. To ensure consistency across reviewers, explicit instructions will be given, and calibration will take place before the extraction process. Two independent reviewers will extract the following data items based on items included in the CHARMS checklist: 1) source of data; 2) participant information; 3) outcome(s) to be predicted; 4) candidate predictors; 5) sample size; 6) Missing data; 7) Model development; 8) Model performance; 9) Model evaluation; 10) Results (final model presented) including model performance; 11) interpretation and discussion. In addition, information will be extracted on the study information (authors, year of publication, study design, journal, contact information, study period, geographical location (area and country), and funding), the assessment of reporting standards using an objective measure if mentioned in the study, and any other relevant information. All relevant text, tables, and figures will be examined for data extraction. Disagreements between the two independent reviewers will be resolved by consensus. We will contact the study authors to request incompletely reported data in included studies. We will conduct analyses using available data if no response is received within 14 days.

### Reporting quality and risk of bias

#### Reporting quality assessment

We will assess the reporting quality of studies against the TRIPOD (Transparent reporting of a multivariable prediction model for individual prognosis or diagnosis) statement, which aims to improve the transparent reporting of prediction modeling studies in all medical settings (31).

The TRIPOD statement provides recommendations for reporting studies on developing, validating, or updating a prediction model. To assess the completeness of reporting amongst each publication, we will utilize the published “TRIPOD Adherence Extraction Form,” which evaluates 22 main items deemed essential in evaluating the transparency of prediction model studies. Each article will only be assessed for items and sub-items that it applies to (development, external validation, or incremental value reporting of prediction models) based on guidance from the adherence form. Each TRIPOD item is given adherence elements to help evaluate an item. The presence or lack of an adherence element within the article will be marked down either as a “Yes” or a “No.” For a TRIPOD item to receive a score, all adherence elements must be present. Overall article’s TRIPOD score can be calculated by summing up adhered TRIPOD items and dividing by the total number of applicable TRIPOD items for that article. Findings on reporting quality from the TRIPOD adherence extraction will be summarised and graphically presented.

#### Risk of bias assessment

We will assess the risk of bias (ROB) of studies by applying PROBAST (prediction model risk of bias assessment tool) (32). PROBAST was developed to assess ROB and applicability concerns of a study that evaluates (e.g., develops or validates) a multivariable diagnostic or prognostic prediction model. Reviewers will assess each study based on the published “PROBAST Assessment Form.” PROBAST is organized into four domains: participants, predictors, outcomes, and analysis. A ROB rating (high, low, or unclear) will be assigned to each domain based on answering signaling questions provided by the PROBAST assessment form. Signaling questions can be answered as yes, probably yes, no, probably no, or no information. Based on ROB results from the four domains, an overall ROB rating and prediction model applicability rating will be given to the prediction model following the recommendations in the PROBAST assessment form. A tabular presentation on PROBAST results for each study will be available. Results will be summarized and graphically presented for each domain.

#### Comments on Methodology

Two independent reviewers will assess the study quality and risk of bias. To improve review consistency, reviewers will be onboarded on general decision rules and given review practice for TRIPOD and PROBAST before conducting an independent review. Disagreements between two reviewers will be resolved by consensus.

We will contact the author if not enough information is available for assessment. We will utilize the available data if the authors do not respond for 14 days. We will present reporting quality and risk of bias assessment in the respective tables.

### Data synthesis

#### Qualitative synthesis

We will provide a qualitative analysis of the studies and their results following standard 4.2 and conduct a qualitative synthesis, chapter 4 of Finding What Works in Health Care: Standards for Systematic Review (33). In addition, we will analyze studies by themes following the primary and secondary outcomes. We will report details of prediction model performance, comparing ML-based and non-ML-based models, the prediction model use case, choice of predictors and outcomes, ML model construct, prediction model reporting standard, and risk of bias.

#### Quantitative synthesis

The studies will likely show significant clinical, methodological and statistical heterogeneity. We will therefore synthesize data quantitatively in appropriate subgroups (prediction of CKD development, CKD diagnosis, and prediction of CKD progression to ESKD), guided by the Cochrane Handbook of Systematic Reviews of Diagnostic Tests Accuracy, Chapter 10: Understanding meta-analysis. Draft version (13 May 2022) (34).

#### Measure of effect size

Performance measures of the ML algorithms will be recorded, including 2×2 confusion matrix, sensitivity, specificity, and AUROC or any other measures utilized by individual studies. The most common measure will be selected for further meta-analysis if feasible.

#### Assessment of heterogeneity

We will assess the clinical heterogeneity of studies based on the ML algorithm use case, the participant characteristics, the predictor choice, and selection. We will assess the methodological heterogeneity based on the ML algorithm construct regarding the data size, ML technique, and performance measures. We will assess statistical heterogeneity using the χ^2^ test and the I^2^ statistic. We will consider an *I*^*2*^ value greater than 50% indicative of substantial heterogeneity.

#### Quantitative Data Synthesis

Suppose sensitivity, specificity, 2×2 confusion matrix results are available for the majority of studies, we will use either a bivariate model or Rutter and Gatsonis HSROC models depending on the consistency of study thresholds to calculate to define a summary point or SROC curve of the algorithm performance estimate combining sensitivity and specificity.

We will undergo exploratory analysis comparing the performance of ML and non-ML techniques. Depending on the number of studies that evaluated both methods, we will either restrict the analysis to these studies or combine all studies that have utilized one of the techniques. We will again use the bivariate model or the Rutter and Gatsonis HSROC model to compare the relative accuracy of both tests.

If the majority of sensitivity, specificity, and 2×2 confusion data are not available, and if AUROC is the most common performance measure, we will utilize methods described in Christodoulou et al. (35) by analyzing pairwise differences in logit AUROCs between ML-based and non-ML based techniques by random effects modeling by DerSimonian and Laird method, either pooled or within subgroups stratified by the risk of bias, study outcomes, ML techniques. Points and the 95% CI will describe the pairwise differences in logit AUROCs.

The meta-analysis will be performed using Review Manager version 5.4.1 (The Cochrane Collaboration, The Nordic Cochrane Centre, Copenhagen, Denmark) (36) and Stata (37).

#### Additional analysis

Further subgroup analyses will be performed to explore possible sources of heterogeneity based on the following: study quality (risk of bias and reporting quality assessment), ML techniques, stages of validation, and dataset (stratified as determined by resulted studies).

We will conduct sensitivity analyses based on study quality, study publication years (stratified by year), study populations, the ML model construct, the ML user case, or any other relevant strata.

### Publication bias

A funnel plot will be constructed to assess the risk of publication bias.

### Confidence in cumulative evidence

Overall evidence quality will be assessed using The Grade of Recommendations of Assessment, Development, and Evaluation (GRADE) guidance for assessing certainty evidence for test accuracy incorporating domains including the risk of bias, indirectness, applicability, inconsistency, imprecision, and publication bias (38). The overall level of evidence: high, moderate, low, very low (one level deducted for each domain with serious concerns/high risk of bias). No factors increased the level.

### Systematic review reporting

As ML in CKD is a rapidly developing field, if the number of studies that meet the eligibility criteria exceeds the capacity to report the study outcomes in one systematic review, the research team will report diagnostic and prognostic tools as separate systematic reviews to ensure a clear and focused reporting and appraisal of study outcomes.

## Discussion

Artificial intelligence and machine learning have significant potential in modern healthcare. Specifically, models developed for the prediction and diagnosis of CKD development and progression can allow for early disease recognition and intervention, which may help to facilitate early disease prevention and diagnosis, assist with early care planning, and allocate resources for the most significant clinical and economic benefit.

As the number of algorithms grows exponentially, the focus should direct toward addressing the barriers to clinical implementation. This review aims to assess the methodological rigor of model development and compare ML-based algorithms’ performance with conventional methods. This will inform clinicians and technical specialists of the current development of ML in CKD care, as well as direct future model development and standardization.

## Data Availability

All relevant data from this study will be made available upon study completion.

## Acknowledgments

None

## Authors’ contributions

Conceptualization: FC, TN, PK, AC, KP

Methodology: FC, TN, PK, AC, KP

Writing – Original Draft Preparation: FC, AC

Writing – Review & Editing: FC, PK, TN, KP

Supervision and guarantor: KP

## References

1. Hill, N. R., Fatoba, S. T., Oke, J. L., Hirst, J. A., O’Callaghan, C. A., Lasserson, D. S., & Hobbs, F. D. (2016). Global Prevalence of Chronic Kidney Disease - A Systematic Review and Meta-Analysis. PloS one, 11(7), e0158765.

2. GBD Chronic Kidney Disease Collaboration (2020). Global, regional, and national burden of chronic kidney disease, 1990-2017: a systematic analysis for the Global Burden of Disease Study 2017. Lancet (London, England), 395(10225), 709–733.

3. Chen, T. K., Knicely, D. H., & Grams, M. E. (2019). Chronic Kidney Disease Diagnosis and Management: A Review. JAMA, 322(13), 1294–1304.

4. Levin, A., Tonelli, M., Bonventre, J., Coresh, J., Donner, J. A., Fogo, A. B., Fox, C. S., Gansevoort, R. T., Heerspink, H., Jardine, M., Kasiske, B., Köttgen, A., Kretzler, M., Levey, A. S., Luyckx, V. A., Mehta, R., Moe, O., Obrador, G., Pannu, N., Parikh, C. R., … ISN Global Kidney Health Summit participants (2017). Global kidney health 2017 and beyond: a roadmap for closing gaps in care, research, and policy. Lancet (London, England), 390(10105), 1888–1917.

5. Liyanage, T., Ninomiya, T., Jha, V., Neal, B., Patrice, H. M., Okpechi, I., Zhao, M. H., Lv, J., Garg, A. X., Knight, J., Rodgers, A., Gallagher, M., Kotwal, S., Cass, A., & Perkovic, V. (2015). Worldwide access to treatment for end-stage kidney disease: a systematic review. Lancet (London, England), 385(9981), 1975–1982.

6. Panch T, Duralde E, Mattie H, Kotecha G, Celi LA, Wright M, et al. (2022) A distributed approach to the regulation of clinical AI. PLOS Digit Health 1(5): e0000040.

7. Busnatu, □., Niculescu, A. G., Bolocan, A., Petrescu, G., Păduraru, D. N., Năstasă, I., Lupu□oru, M., Geantă, M., Andronic, O., Grumezescu, A. M., & Martins, H. (2022). Clinical Applications of Artificial Intelligence-An Updated Overview. Journal of clinical medicine, 11(8), 2265.

8. Choi, R. Y., Coyner, A. S., Kalpathy-Cramer, J., Chiang, M. F., & Campbell, J. P. (2020). Introduction to Machine Learning, Neural Networks, and Deep Learning. Translational vision science & technology, 9(2), 14.

9. Dash, S., Shakyawar, S.K., Sharma, M. et al. Big data in healthcare: management, analysis and future prospects. J Big Data 6, 54 (2019).

10. Kaur, N., Bhattacharya, S., & Butte, A. J. (2021). Big Data in Nephrology. Nature reviews. Nephrology, 17(10), 676–687.

11. Zhang, K., Liu, X., Xu, J., Yuan, J., Cai, W., Chen, T., Wang, K., Gao, Y., Nie, S., Xu, X., Qin, X., Su, Y., Xu, W., Olvera, A., Xue, K., Li, Z., Zhang, M., Zeng, X., Zhang, C. L., Li, O., … Wang, G. (2021). Deep-learning models for the detection and incidence prediction of chronic kidney disease and type 2 diabetes from retinal fundus images. Nature biomedical engineering, 5(6), 533–545. https://doi.org/10.1038/s41551-021-00745-6

12. Kao, H. Y., Chang, C. C., Chang, C. F., Chen, Y. C., Cheewakriangkrai, C., & Tu, Y. L. (2022). Associations between Sex and Risk Factors for Predicting Chronic Kidney Disease. International journal of environmental research and public health, 19(3), 1219.

13. Allen, A., Iqbal, Z., Green-Saxena, A., Hurtado, M., Hoffman, J., Mao, Q., & Das, R. (2022). Prediction of diabetic kidney disease with machine learning algorithms, upon the initial diagnosis of type 2 diabetes mellitus. BMJ open diabetes research & care, 10(1), e002560.

14. Chuah, A., Walters, G., Christiadi, D., Karpe, K., Kennard, A., Singer, R., Talaulikar, G., Ge, W., Suominen, H., Andrews, T. D., & Jiang, S. (2022). Machine Learning Improves Upon Clinicians’ Prediction of End Stage Kidney Disease. Frontiers in medicine, 9, 837232.

15. Lee, K. H., Chu, Y. C., Tsai, M. T., Tseng, W. C., Lin, Y. P., Ou, S. M., & Tarng, D. C. (2022). Artificial Intelligence for Risk Prediction of End-Stage Renal Disease in Sepsis Survivors with Chronic Kidney Disease. Biomedicines, 10(3), 546.

16. Tangri, N., Stevens, L. A., Griffith, J., Tighiouart, H., Djurdjev, O., Naimark, D., Levin, A., & Levey, A. S. (2011). A predictive model for progression of chronic kidney disease to kidney failure. JAMA, 305(15), 1553–1559.

17. Tangri, N., Grams, M. E., Levey, A. S., Coresh, J., Appel, L. J., Astor, B. C., Chodick, G., Collins, A. J., Djurdjev, O., Elley, C. R., Evans, M., Garg, A. X., Hallan, S. I., Inker, L. A., Ito, S., Jee, S. H., Kovesdy, C. P., Kronenberg, F., Heerspink, H. J., Marks, A., … CKD Prognosis Consortium (2016). Multinational Assessment of Accuracy of Equations for Predicting Risk of Kidney Failure: A Meta-analysis. JAMA, 315(2), 164–174.

18. Chan, C. T., Blankestijn, P. J., Dember, L. M., Gallieni, M., Harris, D., Lok, C. E., Mehrotra, R., Stevens, P. E., Wang, A. Y., Cheung, M., Wheeler, D. C., Winkelmayer, W. C., Pollock, C. A., & Conference Participants (2019). Dialysis initiation, modality choice, access, and prescription: conclusions from a Kidney Disease: Improving Global Outcomes (KDIGO) Controversies Conference. Kidney international, 96(1), 37–47.

19. Christodoulou, E., Ma, J., Collins, G. S., Steyerberg, E. W., Verbakel, J. Y., & Van Calster, B. (2019). A systematic review shows no performance benefit of machine learning over logistic regression for clinical prediction models. Journal of clinical epidemiology, 110, 12–22.

20. Nusinovici, S., Tham, Y. C., Chak Yan, M. Y., Wei Ting, D. S., Li, J., Sabanayagam, C., Wong, T. Y., & Cheng, C. Y. (2020). Logistic regression was as good as machine learning for predicting major chronic diseases. Journal of clinical epidemiology, 122, 56–69.

21. Song, X., Liu, X., Liu, F., & Wang, C. (2021). Comparison of machine learning and logistic regression models in predicting acute kidney injury: A systematic review and meta-analysis. International journal of medical informatics, 151, 104484.

22. Collins, G. S., Omar, O., Shanyinde, M., & Yu, L. M. (2013). A systematic review finds prediction models for chronic kidney disease were poorly reported and often developed using inappropriate methods. Journal of clinical epidemiology, 66(3), 268– 277.

23. Ramspek, C. L., de Jong, Y., Dekker, F. W., & van Diepen, M. (2020). Towards the best kidney failure prediction tool: a systematic review and selection aid. Nephrology, dialysis, transplantation : official publication of the European Dialysis and Transplant Association - European Renal Association, 35(9), 1527–1538.

24. Nagendran, M., Chen, Y., Lovejoy, C. A., Gordon, A. C., Komorowski, M., Harvey, H., Topol, E. J., Ioannidis, J., Collins, G. S., & Maruthappu, M. (2020). Artificial intelligence versus clinicians: systematic review of design, reporting standards, and claims of deep learning studies. BMJ (Clinical research ed.), 368, m689.

25. Dhiman, P., Ma, J., Navarro, C. A., Speich, B., Bullock, G., Damen, J. A., Kirtley, S., Hooft, L., Riley, R. D., Van Calster, B., Moons, K., & Collins, G. S. (2021). Reporting of prognostic clinical prediction models based on machine learning methods in oncology needs to be improved. Journal of clinical epidemiology, 138, 60–72.

26. Wynants L, Van Calster B, Collins GS, et al. Prediction models for diagnosis and prognosis of covid-19: systematic review and critical appraisal [published correction appears in BMJ. 2020 Jun 3;369:m2204]. BMJ. 2020;369:m1328.

27. Shamseer, L., Moher, D., Clarke, M., Ghersi, D., Liberati, A., Petticrew, M., Shekelle, P., Stewart, L. A., & PRISMA-P Group (2015). Preferred reporting items for systematic review and meta-analysis protocols (PRISMA-P) 2015: elaboration and explanation. BMJ (Clinical research ed.), 350, g7647.

28. Moons, K. G., de Groot, J. A., Bouwmeester, W., Vergouwe, Y., Mallett, S., Altman, D. G., Reitsma, J. B., & Collins, G. S. (2014). Critical appraisal and data extraction for systematic reviews of prediction modelling studies: the CHARMS checklist. PLoS medicine, 11(10), e1001744.

29. Covidence systematic review software, Veritas Health Innovation, Melbourne, Australia. Available at http://www.covidence.org.

30. Page MJ, McKenzie JE, Bossuyt PM, Boutron I, Hoffmann TC, Mulrow CD, et al. The PRISMA 2020 statement: an updated guideline for reporting systematic reviews. BMJ 2021;372:n71.

31. Collins, G. S., Reitsma, J. B., Altman, D. G., & Moons, K. G. (2015). Transparent reporting of a multivariable prediction model for individual prognosis or diagnosis (TRIPOD): the TRIPOD statement. BMJ (Clinical research ed.), 350, g7594.

32. Wolff, R. F., Moons, K., Riley, R. D., Whiting, P. F., Westwood, M., Collins, G. S., Reitsma, J. B., Kleijnen, J., Mallett, S., & PROBAST Group† (2019). PROBAST: A Tool to Assess the Risk of Bias and Applicability of Prediction Model Studies. Annals of internal medicine, 170(1), 51–58.

33. Standard 4.2 conduct a qualitative synthesis, chapter 4 of Finding What Works in Health Care: Standards for Systematic Reviews

34. Cochrane Handbook of Systematic Reviews of Diagnostic Tests Accuracy, Chapter 10: Understanding meta-analysis. Draft version (13 May 2022)

35. Christodoulou, E., Ma, J., Collins, G. S., Steyerberg, E. W., Verbakel, J. Y., & Van Calster, B. (2019). A systematic review shows no performance benefit of machine learning over logistic regression for clinical prediction models. Journal of clinical epidemiology, 110, 12–22.

36. Review Manager 5 (RevMan 5). 5.3 ed. Copenhagen: Nordic Cochrane Centre, The Cochrane Collaboration; 2014.

37. StataCorp. 2021. Stata Statistical Software: Release 17. College Station, TX: StataCorp LLC.

38. Yang B, Mustafa RA, Bossuyt PM, et al. GRADE Guidance: 31. Assessing the certainty across a body of evidence for comparative test accuracy. J Clin Epidemiol. 2021;136:146–156.

