## Additional File for "Prediction and diagnosis of chronic kidney disease development and progression using machine-learning: protocol for a systematic review and meta-analysis of reporting standards and model performance"

**Search Strategy**

**Search Date: 14/09/2022**

**Search concept**

- Chronic Kidney disease
- Machine Learning

| Set 1 | **Pubmed** | Results |
| --- | --- | --- |
| 1 | "renal insufficiency, chronic"[MeSH Terms] OR "kidney failure, chronic"[MeSH Terms] OR ((chronic[tiab] OR (end[tiab] AND stage*[tiab]) OR endstage*[tiab] OR "end-stage"[tiab]) AND (((kidney[tiab] OR renal[tiab]) AND (fail*[tiab] OR disease*[tiab])) OR nephropath*[tiab])) OR "chronic kidney failure*"[tiab] OR "chronic renal failure*"[tiab] OR "chronic kidney disease*"[tiab] OR "end-stage kidney disease*"[tiab] OR "end stage kidney disease*"[tiab] OR "endstage kidney disease*"[tiab] OR "end-stage renal disease*"[tiab] OR "end stage renal disease*"[tiab] OR "endstage renal disease*"[tiab] OR CRF[tiab] OR CRD[tiab] OR CKF[tiab] OR CKD[tiab] OR ESRF[tiab] OR ESRD[tiab] OR ESKF[tiab] OR ESKD[tiab] OR ((kidney[tiab] OR renal[tiab]) AND insufficien*[tiab]) OR "kidney insufficien*"[tiab] OR "renal insufficien*"[tiab] | 265,539 |
| 2 | "artificial intelligence"[MeSH Terms] OR ((artificial[tiab] OR machine*[tiab]) AND (intelligen*[tiab] OR reason*[tiab] OR aid[tiab] OR aids[tiab] OR aided[tiab] OR diagnos*[tiab] OR system*[tiab] OR (knowledge[tiab] AND (acquisition[tiab] OR acquire*[tiab] OR representation*[tiab])) OR interpretation*[tiab] OR recogni*[tiab] OR classif*[tiab])) OR "artificial intelligence"[tiab] OR "artificial intelligent"[tiab] OR "artificial reasoning"[tiab] OR "machine learning"[MeSH Terms] OR "deep learning"[MeSH Terms] OR ((machine[tiab] OR transfer[tiab] OR deep[tiab] OR hierarch*[tiab] OR supervised[tiab] OR unsupervised[tiab] OR "un-supervised"[tiab] OR reinforcement[tiab]) AND learning[tiab]) OR "machine learning"[tiab] OR "transfer learning"[tiab] OR "deep learning"[tiab] OR "hierarchical learning"[tiab] OR "supervised learning"[tiab] OR "unsupervised learning"[tiab] OR "un-supervised learning"[tiab] OR "reinforcement learning"[tiab] OR "neural networks, computer"[MeSH Terms] OR (neural[tiab] AND network*[tiab]) OR "neural network*"[tiab] OR perceptron*[tiab] OR (connectionist[tiab] AND model*[tiab]) OR "connectionist model"[tiab] OR "connectionist models"[tiab] OR "support vector machine"[MeSH Terms] OR (support[tiab] AND vector[tiab] AND (machine*[tiab] OR network*[tiab])) OR "support vector machine"[tiab] OR "support vector network"[tiab] OR (data[tiab] AND mining[tiab]) OR "data mining"[tiab] | 414,227 |
| 3 | #1 AND #2 | 2026 |
| 4 | Animals[Mesh Terms] NOT humans[Mesh Terms] | 5,041,752 |
| 5 | #3 NOT #4 | **1,918** |

| Set 2 | **Embase** | Results |
| --- | --- | --- |
| 1 | exp chronic kidney failure/ OR exp end stage renal disease/ OR ('chronic'.ti,ab OR ('end’ .ti,ab AND 'stage*' .ti,ab) OR 'endstage*' .ti,ab OR 'end-stage' .ti,ab) AND ((‘kidney’ .ti,ab OR ‘renal’ .ti,ab) AND (‘fail*’ .ti,ab OR ‘disease*’ .ti,ab)) OR nephropath* .ti,ab) OR 'chronic kidney failure*' .ti,ab OR 'chronic renal failure*' .ti,ab OR 'chronic kidney disease*' .ti,ab OR 'end-stage kidney disease*' .ti,ab OR 'end stage kidney disease*' .ti,ab OR 'endstage kidney disease*' .ti,ab OR 'end-stage renal disease*' .ti,ab OR 'end stage renal disease*' .ti,ab OR 'endstage renal disease*' .ti,ab OR 'CRF' .ti,ab OR 'CRD' .ti,ab OR 'CKF' .ti,ab OR 'CKD' .ti,ab OR 'ESRF' .ti,ab OR 'ESRD' .ti,ab OR 'ESKF' .ti,ab OR 'ESKD' .ti,ab OR (('kidney' .ti,ab OR 'renal' .ti,ab) AND 'insufficien*' .ti,ab) OR 'kidney insufficien*' .ti,ab OR 'renal insufficien*' .ti,ab | 399,235 |
| 2 | exp artificial intelligence/ OR (('artificial' .ti,ab OR 'machine*' .ti,ab) AND ('intelligen*' .ti,ab OR 'reason*' .ti,ab OR 'aid*' .ti,ab OR 'diagnos*' .ti,ab OR 'system*' .ti,ab OR ('knowledge' .ti,ab AND ('acquisition' .ti,ab OR 'acquire*' .ti,ab OR 'representation*' .ti,ab)) OR 'interpretation*' .ti,ab OR 'recogni*' .ti,ab OR 'classif*' .ti,ab)) OR 'artificial intelligence' .ti,ab OR 'artificial intelligent' .ti,ab OR 'artificial reasoning' .ti,ab OR exp machine learning/ OR exp deep learning/ OR (('machine' .ti,ab OR 'transfer' .ti,ab OR 'deep' .ti,ab OR 'hierarch*' .ti,ab OR 'supervised' .ti,ab OR 'unsupervised' .ti,ab OR 'un-supervised' .ti,ab OR 'reinforcement' .ti,ab) AND 'learning' .ti,ab) OR 'machine learning' .ti,ab OR 'transfer learning' .ti,ab OR 'deep learning' .ti,ab OR 'hierarchical learning' .ti,ab OR 'supervised learning' .ti,ab OR 'unsupervised learning' .ti,ab OR 'un-supervised learning' .ti,ab OR 'reinforcement learning' .ti,ab OR exp artificial neural network/ OR ('neural' .ti,ab AND 'network*' .ti,ab) OR ' neural network*' .ti,ab OR 'perceptron*' .ti,ab OR ('connectionist' .ti,ab AND 'model*' .ti,ab) OR 'connectionist model' .ti,ab OR 'connectionist models' .ti,ab OR exp support vector machine/ OR ('support' .ti,ab AND 'vector' .ti,ab AND ('machine*' .ti,ab OR 'network*' .ti,ab)) OR 'support vector machine' .ti,ab OR 'support vector network' .ti,ab OR ('data' .ti,ab AND 'mining' .ti,ab) OR 'data mining' .ti,ab | 592,402 |
| 3 | #1 AND #2 | 3,696 |
| 4 | [animals]/lim NOT [humans]/lim | 5,782,519 |
| 5 | #3 NOT #4 | **3,482** |

| Set 3 | **Web of Science** | Results |
| --- | --- | --- |
| 1 | TS=(((chronic OR (end AND stage*) OR endstage* OR "end-stage") AND (((kidney OR renal) AND (fail* OR disease*)) OR nephropath*)) OR "chronic kidney failure*" OR "chronic renal failure*" OR "chronic kidney disease*" OR "end-stage kidney disease*" OR "end stage kidney disease*" OR "endstage kidney disease*" OR "end-stage renal disease*" OR "end stage renal disease*" OR "endstage renal disease*" OR CRF OR CRD OR CKF OR CKD OR ESRF OR ESRD OR ESKF OR ESKD OR ((kidney OR renal) AND insufficien*) OR "kidney insufficien*" OR "renal insufficien*") | 250,490 |
| 2 | TS=(((artificial OR machine*) AND (intelligen* OR reason*OR aid OR aids OR aided OR diagnos* OR system* OR (knowledge AND (acquisition OR acquire* OR representation*)) OR interpretation* OR recogni* OR classif*)) OR “artificial intelligence” OR “artificial intelligent” OR “artificial reasoning” OR ((machine OR transfer OR deep OR hierarch* OR supervised OR unsupervised OR “un-supervised” OR reinforcement) AND learning) OR “machine learning” OR “transfer learning” OR “deep learning” OR “hierarchical learning” OR “supervised learning” OR “unsupervised learning” OR “un-supervised learning” OR “reinforcement learning” OR (neural AND network*) OR “neural network*” OR perceptron* OR (connectionist AND model*) OR “connectionist model” OR “connectionist models” OR (support AND vector AND (machine* OR network*)) OR “support vector machine” OR “support vector network” OR (data AND mining) OR “data mining”) | 1,579,559 |
| 3 | #1 AND #2 | 3,843 |
| 4 | ALL=(animal NOT human) | 1,511,739 |
| 5 | #3 NOT #4 | **3,783** |

| Set 4 | **CENTRAL** | Results |
| --- | --- | --- |
| 1 | [mh "renal insufficiency, chronic"] OR [mh "kidney failure, chronic"] OR (((chronic):ti,ab,kw OR ((end):ti,ab,kw AND (stage*):ti,ab,kw) OR (endstage*):ti,ab,kw OR ("end-stage"):ti,ab,kw) AND ((((kidney):ti,ab,kw OR (renal):ti,ab,kw) AND ((fail*):ti,ab,kw OR (disease*):ti,ab,kw)) OR (nephropath*):ti,ab,kw)) OR ("chronic kidney failure*"):ti,ab,kw OR ("chronic renal failure*"):ti,ab,kw OR ("chronic kidney disease*"):ti,ab,kw OR ("end-stage kidney disease*"):ti,ab,kw OR ("end stage kidney disease*"):ti,ab,kw OR ("endstage kidney disease*"):ti,ab,kw OR ("end-stage renal disease*"):ti,ab,kw OR ("end stage renal disease*"):ti,ab,kw OR ("endstage renal disease*"):ti,ab,kw OR (CRF):ti,ab,kw OR (CRD):ti,ab,kw OR (CKF):ti,ab,kw OR (CKD):ti,ab,kw OR (ESRF):ti,ab,kw OR (ESRD):ti,ab,kw OR (ESKF):ti,ab,kw OR (ESKD):ti,ab,kw OR (((kidney):ti,ab,kw OR (renal):ti,ab,kw) AND (insufficien*):ti,ab,kw) OR ("kidney insufficien*"):ti,ab,kw OR ("renal insufficien*"):ti,ab,kw | 31,469 |
| 2 | [mh "artificial intelligence"] OR [mh "machine learning"] OR [mh "deep learning"] OR [mh "support vector machine"] OR [mh "neural networks, computer"] OR (((artificial):ti,ab,kw OR (machine*):ti,ab,kw) AND ((intelligen*):ti,ab,kw OR (reason*):ti,ab,kw OR (aid):ti,ab,kw OR (aids):ti,ab,kw OR (aided):ti,ab,kw OR (diagnos*):ti,ab,kw OR (system*):ti,ab,kw OR ((knowledge):ti,ab,kw AND ((acquisition):ti,ab,kw OR (acquire*):ti,ab,kw OR (representation*):ti,ab,kw)) OR (interpretation*):ti,ab,kw OR (recogni*):ti,ab,kw OR (classif*):ti,ab,kw)) OR ("artificial intelligence"):ti,ab,kw OR ("artificial intelligent"):ti,ab,kw OR ("artificial reasoning"):ti,ab,kw OR (((machine):ti,ab,kw OR (transfer):ti,ab,kw OR (deep):ti,ab,kw OR (hierarch*):ti,ab,kw OR (supervised):ti,ab,kw OR (unsupervised):ti,ab,kw OR ("un-supervised"):ti,ab,kw OR (reinforcement):ti,ab,kw) AND (learning):ti,ab,kw) OR ("machine learning"):ti,ab,kw OR ("transfer learning"):ti,ab,kw OR ("deep learning"):ti,ab,kw OR ("hierarchical learning"):ti,ab,kw OR ("supervised learning"):ti,ab,kw OR ("unsupervised learning"):ti,ab,kw OR ("un-supervised learning"):ti,ab,kw OR ("reinforcement learning"):ti,ab,kw OR ((neural):ti,ab,kw AND (network*):ti,ab,kw) OR ("neural network*"):ti,ab,kw OR (perceptron*):ti,ab,kw OR ((connectionist):ti,ab,kw AND (model*):ti,ab,kw) OR ("connectionist model"):ti,ab,kw OR ("connectionist models"):ti,ab,kw OR ((support):ti,ab,kw AND (vector):ti,ab,kw AND ((machine*):ti,ab,kw OR (network*):ti,ab,kw)) OR ("support vector machine"):ti,ab,kw OR ("support vector network"):ti,ab,kw OR ((data):ti,ab,kw AND (mining):ti,ab,kw) OR ("data mining"):ti,ab,kw | 19,738 |
| 3 | #1 AND #2 | **429** |

| Set 5 | **IEEE Xplore** | Results |
| --- | --- | --- |
| 1 | ((((chronic OR (end AND stage*) OR endstage* OR "end-stage") AND (((kidney OR renal) AND (fail* OR disease OR diseases OR diseased)) OR nephropath*)) OR "chronic kidney failure" OR "chronic renal failure" OR "chronic kidney disease" OR "chronic kidney diseases" OR "end-stage kidney disease" OR "end stage kidney disease" OR "endstage kidney disease" OR "end-stage renal disease" OR "end stage renal disease" OR "endstage renal disease" OR CRF OR CRD OR CKF OR CKD OR ESRF OR ESRD OR ESKF OR ESKD OR ((kidney OR renal) AND insufficien*) OR "kidney insufficien*" OR "renal insufficien*")) | 3955 |
| 2 | ((((artificial OR machine OR machines OR machinery) AND (intelligen* OR reason* OR aid OR aids OR aided OR diagnos* OR system* OR (knowledge AND (acquisition OR acquire OR representation OR representational)) OR interpretation* OR recogni* OR classif*)) OR "artificial intelligence" OR "artificial intelligent" OR "artificial reasoning" OR ((machine OR transfer OR deep OR hierarch* OR supervised OR unsupervised OR "un-supervised" OR reinforcement) AND learning) OR "machine learning" OR "transfer learning" OR "deep learning" OR "hierarchical learning" OR "supervised learning" OR "unsupervised learning" OR "un-supervised learning" OR "reinforcement learning" OR (neural AND (network OR networks)) OR "neural network" OR "neural networks" OR perceptron OR perceptrons OR (connectionist AND model) OR "connectionist model" OR "connectionist models" OR (support AND vector AND (machine OR machines OR machinery OR network)) OR "support vector machine" OR "support vector network" OR (data AND mining) OR "data mining")) | 936,877 |
| 3 | 1 AND 2 | **1870** |
